# Neurologic sequalae of COVID-19 are determined by immunologic imprinting from previous Coronaviruses

**DOI:** 10.1101/2022.11.07.22282030

**Authors:** Marianna Spatola, Nadège Nziza, Wonyeong Jung, Yixiang Deng, Dansu Yuan, Alessandro Dinoto, Silvia Bozzetti, Vanessa Chiodega, Sergio Ferrari, Douglas A Lauffenburger, Sara Mariotto, Galit Alter

**Affiliations:** Ragon Institute of MGH, MIT and Harvard Medical School, Cambridge, MA, USA; Massachusetts Institute of Technology, Cambridge, MA, USA; Neurology Unit, Department of Neuroscience, Biomedicine and Movement Sciences, University of Verona, Verona, Italy; Department of Neurology/Stroke Unit, San Maurizio hospital, Bolzano, Italy

**Keywords:** SARS-CoV-2, Fc receptor, antibody-mediated complement activation, antibody-mediated phagocytosis, PASC, long-COVID

## Abstract

Coronavirus disease 2019 (COVID-19), which is caused by the severe acute respiratory syndrome coronavirus 2 (SARS-CoV-2), remains a global public health emergency. Although SARS-CoV-2 is primarily a respiratory pathogen, extra-respiratory organs, including the central nervous system (CNS), can also be affected. Neurologic symptoms have been observed not only during acute SARS-CoV-2 infection, but also at distance from respiratory disease, also known as long-COVID or neurological post-acute sequelae of COVID-19 (neuroPASC). The pathogenesis of neuroPASC is not well understood, but hypotheses include SARS-CoV-2-induced immune dysfunctions, hormonal dysregulations, and persistence of SARS-CoV-2 reservoirs. In this study, we used a high throughput systems serology approach to dissect the humoral response to SARS-CoV-2 (and other common Coronaviruses - 229E, HKU1, NL63, OC43) in the serum and cerebrospinal fluid (CSF) from 112 infected individuals who developed or did not develop neuroPASC. Unique SARS-CoV-2 humoral profiles were observed in the CSF of neuroPASC. All antibody isotypes (IgA, IgM, IgA) and subclasses (IgA1-2; IgG1-4) were detected in serum, whereas CSF was characterized by focused IgG1 (and absence of IgM). These data argue in favor of compartmentalized brain-specific responses against SARS-CoV-2 through selective transfer of antibodies from the serum to the CSF across the blood-brain-barrier, rather than intrathecal synthesis, where more diversity in antibody classes/subclasses would be expected. Moreover, compared to individuals who did not develop post-acute neurological complications following infection (n=94), those with neuroPASC (n=18) exhibited attenuated systemic antibody responses against SARS-CoV-2, characterized by decreased capacity to activate antibody-dependent complement deposition (ADCD), NK cell activation (ADNKA) and to bind Fcγ receptors. However, surprisingly, neuroPASC showed significantly expanded antibody responses to other common Coronaviruses, including 229E, HKU1, NL63, and OC43. This biased humoral activation across coronaviruses was particularly enriched in neuroPASC individuals with poor outcome, suggesting an *original antigenic sin* (or immunologic imprinting), where pre-existing immune responses against related viruses shape the response to current infection, as a key prognostic marker of neuroPASC disease. Overall, these findings point to a pathogenic role for compromised anti-SARS-CoV-2 responses in the CSF, likely resulting in incomplete virus clearance from the brain and persistent neuroinflammation, in the development of post-acute neurologic complications of SARS-CoV-2 infection.

## INTRODUCTION

Coronavirus disease 2019 (COVID-19), which is caused by the severe acute respiratory syndrome coronavirus 2 (SARS-CoV-2),^1,2^ remains a global public health emergency. Although SARS-CoV-2 is primarily a respiratory pathogen, extra-respiratory organs, including the central nervous system (CNS), can also be affected.^3-6^ Despite analyses of cerebrospinal fluid (CSF) from living individuals have found limited to no evidence of active viral replication in the CSF,^7^ neurologic symptoms can develop following SARS-CoV-2 infection, both acutely and at distance from initial respiratory disease, as part of neurological post-acute sequelae of COVID-19 (neuroPASC).^8,9^ These symptoms include impaired concentration, headache, confusion, cognitive disturbances or even encephalitis, persisting, in some cases, for several months after recovery from infection.^10-12^ The pathophysiological mechanisms of this complication are not well understood, but hypotheses include COVID-19 driven immune dysfunctions,^13^ impaired vagus nerve signaling,^10,13^ hormonal dysregulations,^8^ persistence of SARS-CoV-2 reservoirs,^14^ auto-antibody evolution,^13,15^ interactions between SARS-CoV-2 and the host virome or microbiome,^16^ or reactivation of neurotrophic infections.^17,18^

Emerging data point to incomplete clearance of SARS-CoV-2 infection as a marker of post-COVID-19 persisting symptoms.^19^ Whether this prolonged viral replication is related to formation of a viral reservoir or related to an incomplete immune response unable to eliminate the virus, remains unclear. Given our emerging appreciation of the importance of antibodies, both as markers of previous pathogen exposure and as potential contributors to local pathology, in this study, we aimed to determine whether alterations in the humoral immune response to SARS-CoV-2 in the serum and CSF might be found in individuals with neuroPASC. Using a high throughput systems serology approach, we identified distinctive humoral signatures across individuals with and without neuroPASC. Individuals with neuroPASC were characterized by a diminished systemic response to SARS-CoV-2, in contrast with an expanded systemic response to common Coronaviruses. These features were also observed in CSF and were particularly enriched in individuals with poor outcome. Moreover, neuroPASC was marked by a distinct CSF and systemic humoral response, characterized by a highly compartmentalized brain-specific antibody signature, pointing to limited evidence of intrathecal SARS-CoV-2 antibody production. Together, these data point to compromised peripheral SARS-CoV-2 immunity, biased towards enhanced recognition of common Coronaviruses, as a signature of imprinting or original antigenic sin, and prognostic marker in neuroPASC.

## MATERIALS AND METHODS

### Study population and biological samples

We recruited individuals with SARS-CoV-2 infection who developed or did not develop neurological post-acute complications of COVID-19 (neuroPASC versus no PASC, respectively). Participants were identified prospectively during the first wave of COVID-19 between March 2020 and August 2020 at the University Hospital of Verona, Italy. NeuroPASC patients were included in the study if they fulfilled the following criteria: 1) new neurological manifestations or significant change in their pre-existing neurological status without alternative explanation/cause; 2) history of COVID-19 symptoms with PCR-proven SARS-CoV-2 infection, or 3) no history of COVID-19 symptoms but previous SARS-CoV-2 infection proven by either PCR on nasal swab (in case they were tested despite being asymptomatic) or by positive SARS-CoV-2 IgG serology. As controls (no PASC), we selected health care professionals with PCR-proven SARS-CoV-2 infection, that were tested by nasal swab as part of the routine weekly SARS-CoV-2 surveillance. Of note, none of the participants received any SARS-CoV-2 vaccination, which was not yet available at the time of the study. Also, all included individuals (both neuroPASC and no PASC) were re-tested at the University Hospital of Verona to confirm positive SARS-CoV-2 IgG serology by ELISA. Serum samples were obtained at onset of neurological symptoms for patients with neuroPASC, and at 2-4 months follow up for controls (no PASC). CSF was available from neuroPASC patients only, if required for diagnostic assessment, and was obtained within 2 weeks from neurologic onset.

### Outcome measurements

Individuals with neuroPASC were followed up clinically by the referring physician. Outcome was assessed at the last follow-up (at least 6 months after neurological onset) by using a modified Rankin Scale (mRS).^20^ Individuals were considered to have *good* outcome if mRS was <2, or *poor* outcome if mRS was ≥2.

### Antigens

The antibody responses were assessed against 5 different SARS-CoV-2 antigens, including Spike WT (Sino Biological 40589-V08H4), RBD WT (Sino Biological 40592-V08H), S1 WT (Sino Biological 40591-V08H), S2 WT (Sino Biological 40590-V08B) and Nucleocapsid (Sino Biological 40588-V08B), as well as 4 non-SARS-CoV-2 coronaviruses, including HCoV-OC43 (Sino Biological 40607-V08B), -HKU1 (Sino Biological 40606-V08B), -NL63 (Sino Biological 40604-V08B), -229E (Sino Biological 40605-V08B). Non-coronavirus control antigens included herpes simplex virus 1 (HSV1; ImmuneTech IT-005-055p), Epstein-Bar virus (EBV; ImmuneTech IT-005-035p) and Influenza virus A (ImmuneTech IT-003-SW12p and ImmuneTech IT-003-001p).

### Antibody subclass, isotype and FcR binding

Antigen-specific antibody subclass and isotype levels, in addition to Fc receptors (FcRs) binding were measured in serum and CSF samples using a customized multiplexed Luminex assay, as previously reported.^21^ Briefly, each antigen was coupled to different magnetic Luminex beads by carbodiimide-NHS coupling. Serum (dilution 1:100) or CSF (dilution 1:10) samples were then added to the beads to form immune complexes. After a 2 hour incubation at room temperature, immune complexes were washed, and subclasses and isotypes were detected using phycoerythrin (PE)-conjugated mouse anti-human IgG1, IgG2, IgG3, IgG4, IgA1, IgA2 or IgM (Southern Biotech) at 1.3 µg/ml. Concerning FcRs, they were purchased from Duke Human Vaccine Institute, and binding capacity to Avi-tagged FcγR2A, FcγR2B, FcγR3A, FcγR3B, FcαR and neonatal FcR (FcRn) was assessed using PE-streptavidin (Agilent Technologies) that was coupled to the different FcRs. After 1 hour of incubation with subclasses/isotypes or FcRs, immune complexes were washed, and median fluorescence intensity (MFI) was determined on an iQue analyzer (Intellicyt).

### Antibody-dependent cellular phagocytosis (ADCP)

For ADCP, THP-1 monocyte phagocytosis was assessed as previously described.^22^ Antigens were biotinylated and coupled to yellow-green fluorescent neutravidin microspheres (Thermo Fisher F8776), followed with an incubation with either serum (dilution 1:100) or CSF (dilution 1:10). After washing the immune complexes, THP-1 monocytes (0.25 M cells per well) were added and incubated for 16 h at 37°C. Cells were then fixed with 4% paraformaldehyde and analyzed by flow cytometry on an iQue analyzer (Intellicyt). Phagocytosis score was calculated as the (percentage of microsphere-positive cells) × (MFI of microsphere-positive cells) divided by 100 000.

### Antibody-dependent neutrophil phagocytosis (ADNP)

Similarly to ADCP, ADNP was performed using a microsphere-based phagocytic assay, as described previously.^19^ Briefly, after the formation of immune complexes composed of antigen-coupled neutravidin microspheres (Thermo Fisher F8776) and antibodies from serum (dilution 1:100) or CSF (dilution 1:10), human neutrophils obtained from healthy donors’ blood were added and incubated for 1 hour at 37 °C. Neutrophils were surface-stained with anti-CD66b Pac blue antibody (BioLegend 305112) then fixed with 4% paraformaldehyde and analyzed by flow cytometry on an iQue analyzer (Intellicyt). Phagocytosis score was calculated as the (percentage of microsphere-positive CD66b cells) × (MFI of microsphere-positive cells) divided by 100 000.

### Antibody-dependent complement deposition (ADCD)

ADCD was performed as previously described.^23^ In brief, antigens were biotinylated and coupled to red fluorescent neutravidin microspheres (Thermo Fisher F8775), followed by an incubation with serum (dilution 1:40) or CSF (dilution 1:5) samples for 2 hours at 37 °C. Immune complexes were then washed and incubated with complement factors from guinea pig (Cedarlane CL4051) during 20 minutes at 37 °C. To stop the complement reaction, beads were washed with 15 mM EDTA. The deposition of complement was detected by 1:100 diluted fluorescein-conjugated goat IgG to guinea pig complement C3b (MP Biomed 855385), and relative C3 deposition was quantified by flow cytometry on an iQue analyzer (Intellicyt).

### Antibody-dependent natural killer cell activation (ADNKA)

An enzyme-linked immunosorbent assay (ELISA) was performed to measure ADNKA, as previously described.^24^ Briefly, ELISA plates were coated with antigens by incubation overnight at 4 °C, then serum (dilution 1:100) or CSF (dilution 1:10) samples were added for a 2-hour incubation at 37 °C. The day before adding the samples, NK cells were isolated from buffy coats, generated from healthy donors, using RosetteSep NK enrichment kit (Stem Cell Technologies 15065), and were incubated overnight at 37 °C with 1 ng/ml of interleukin-15 (IL-15). After the 2-hour sample incubation, NK cells were added to the ELISA plates and stained with anti-CD107a (BD 555802) and treated with a protein-transport inhibitor (BD Bioscience 554724), and with brefeldin A (Sigma B7651). After a 5-hour incubation at 37 °C, surface cell staining was performed with anti-CD3 (BD 558117) and anti-CD56 (BD 557747) during a 20-minute incubation at room temperature. NK cells were then fixed using Perm A and Perm B (Thermo Fisher GAS001S100) to allow intracellular staining with anti-IFNγ (BD 340449), and anti-MIP1β (BD 550078). NK cells were then analyzed by flow cytometry on an iQue analyzer (Intellicyt) and defined as CD3^−^CD56^+^ cells, and activation was identified as CD107^+^IFNγ^+^MIP1β ^+^.

### Statistical and computational analysis

Univariate comparative analyses of antibody features between groups were performed using non-parametric tests (Mann-Whitney, Kruskal-Wallis, Friedman test) and Fisher exact tests for frequency comparisons between NeuroPASC and no PASC, and between serum and CSF responses. Analyses were carried out using GraphPad Prism with corrections for multiple comparisons. Spearman correlation coefficients were calculated, and p-values were obtained for two-tailed tests.

Multivariate analyses were performed with R (version 4.1.0). Luminex data were log10-transformed. All data were z-scored. Missing values were imputed using R package ‘DMwR’. A partial least square discriminant analysis (PLSDA) with Least Absolute Shrinkage Selection Operator (LASSO) was used to select antibody features that contributed most to discriminate between neuroPASC and no PASC, and between serum and CSF (multilevel PLSDA using paired samples).^25^ Antibody features were ranked according to their Variable Importance in Projection (VIP) scores, for which higher values indicate higher contribution to the model, and their first latent variable loadings (LV1) were shown. Ten-fold cross validation was performed and median p-values were reported. The model was validated by comparing it to a null model, for which the modelling approach was repeated 100x with random shuffling of labels or permutated labels. P-values were obtained from the tail probability of the accuracy distribution of the control model.

### Study approval

All included subjects have given their informed consent to be included in the study. The study has been approved by the local Ethical Committee of the University of Verona.

## RESULTS

### NeuroPASC individuals have decreased systemic SARS-CoV-2 antibody titers, FcγR binding and antibody Fc-functions

A number of serological, transcriptional, and cellular markers^26-28^ have begun to emerge associated with persistent symptoms following the resolution of SARS-CoV-2 infection. However, the markers vary widely, likely due to the strikingly diverse clinical manifestations and pathologies associated with different post-acute sequelae of COVID-19 (PASC), including respiratory, vascular, rheumatological, and neurological complications. However, the identification of specific markers of PASC associated with each clinical phenotype may have prognostic implications and provide critical insights into the etiologic mechanisms that may be targeted to prevent these post-infectious complications. To begin to address whether unique serological markers could be defined in a highly selected group of individuals with neuroPASC, we identified 18 individuals with neuroPASC and 94 without PASC (no PASC). Serum samples were available for antibody studies from all neuroPASC and no PASC individuals. In addition, CSF samples were available from 16 of the neuroPASC individuals for compartment-specific immune profiling.

Clinical manifestations, demographics, results from ancillary tests and outcome of patients with neuroPASC are shown in **Table 1**. Compared to individuals who did not develop PASC, individuals with neuroPASC had similar demographic features (median age 65 vs 66.5 years, respectively, p=0.55; females 33% vs 44%, p=0.52).

**Table 1.** Clinical characteristics of neuroPASC patients.

| **Pt#** | **COVID-19 symptoms** | **Delay from start of pandemic to neurological onset (days)\*; neuroPASC symptoms** | **MRI** | **CSF analysis** | **Treatment** | **Outcome; mRS** |
| --- | --- | --- | --- | --- | --- | --- |
| **1** | Fever, cough | 27; ENC, headache, akinetic mutism, seizures, oculomotor paralysis, dysphagia | Bilateral hippocampal FLAIR hyperintensities | 16 WBC | Dexamethasone; levetiracetam, diazepam, lacosamide, HCQ, gancyclovir, antibiotics | Death; 6 |
| **2** | None | 31; ENC, confusion, anxiety, agitation, psychomotor slowing | Normal | <5 WBC | Iv steroids, HCQ, gancyclovir | Complete recovery; 0 |
| **3** | Dyspnea, fever | 47; ENC | Bilateral hippocampal FLAIR hyperintensities | <5 WBC; elevated protein (77) | IvIg | Improved; N/A |
| **4** | None | 31; ENC, gait disturbance | Multiple subcortical and brainstem FLAIR hyperintensities suggestive of encephalitis | <5 WBC; elevated protein (58) | IvIg | Markedly improved; N/A |
| 5 | None | 41; ENC, delirium | Normal | Normal WBC,<br>normal proteins | Sertraline, olanzapine | Stable; N/A |
| 6 | Dyspnea,<br>cough, fever | 49; ENC | Cortical atrophy | No LP | Levetiracetam, lacosamide<br>gancyclovir, antibiotic, antimycotic | Death; 6 |
| 7 | None | 67; ENC, confusion, seizures,<br>agitation, short-term memory<br>loss | Non-specific areas of<br>gliosis | <5 WBC, elevated<br>protein (50) | levetiracetam | Partial recovery; 1 |
| 8 | None | N/A; Transverse myelitis with<br>T6 sensory level | Spinal cord FLAIR<br>hyperintensity (T6) | 20 WBC, elevated<br>protein (50) | N/A | Partial recovery; 4 |
| 9 | Anosmia,<br>ageusia | 41; ENC, bilateral visual<br>impairment associated with<br>sensory deficit on R leg | Normal | 22 WBC | Iv steroids, IvIg | Improved; N/A |
| 10 | Pneumonia | 92; ENC, confabulation,<br>complete oculomotor palsy,<br>seizures, rigidity | Unilateral hippocampal<br>FLAIR hyperintensity | <5 WBC | IvIg, antiepileptics | N/A |
| 11 | N/A | 32; unilateral decreased visual<br>acuity | Optic nerve and multiple<br>subcortical FLAIR<br>hyperintensities | <5 WBC,<br>increased IgG<br>index | Iv steroids | Partial recovery; 1 |
| 12 | N/A | 55; unilateral decreased visual | N/A | No LP | Iv steroids with oral steroids | Partial recovery; 1 |

|  |  | acuity |  |  | tapering |  |
| --- | --- | --- | --- | --- | --- | --- |
| **13** | None | 136; ENC, confusion, ataxia, motor hemisindrome (stroke), seizures and status epilepticus | Multiple areas of altered signal in the R fronto-temporal and L cerebellar regions with diffusion restriction. | No LP | N/A | N/A |
| **14** | Fever | 145; severe worsening of motor sequelae of transverse myelitis | Sequelae of known transverse myelitis | <5 WBC, normal proteins | N/A | Stable; 5 |
| **15** | None | 93; ENC, personality change, psychosis, decreased level of consciousness | Hippocampal FLAIR hyperintensity cortical atrophy | <5 WBC, normal proteins | None | Stable; 5 |
| **16** | N/A | 137; ENC, confusion, seizures | N/A | No LP | N/A | N/A |
| **17** | None | 163; ENC, status epilepticus | Normal | <5 WBC, normal proteins | Ig | Improved; N/A |
| **18** | None | N/A, transverse myelitis | N/A | 10 WBC, normal proteins | N/A | Stable; 3 |

\*As the delay from SARS-CoV-2 infection to neuroPASC was unknown for many patients, we reported the delay from the start of the pandemic to onset of neurological symptoms as the longest time from possible infection to neurological onset
Legend: ENC = encephalopathy; HCQ = Hydroxychloroquine; IvIg = intravenous immunoglobulins; LP= lumbar puncture; WBC = white blood cells/mm<sup>3</sup> of cerebrospinal fluid (>5 abnormal)

Overall antibody profiling pointed to 2 major observations: 1) every individual possessed a unique SARS-CoV-2 specific antibody profile and 2) individuals with neuroPASC exhibited overall lower systemic antibody responses to various SARS-CoV-2 antigens (Spike, S1, S2, RBD, NC) compared to individuals that did not develop neuroPASC (**Fig 1A**). Specifically, individuals with neuroPASC elicited lower Spike-specific IgG and IgA (but not IgM) titers, and lower Spike-specific FcγR binding capacity and antibody Fc-functions (**Fig 1B-D**). In particular, Spike-specific antibody IgG1 (and IgG2), antibody dependent complement deposition (ADCD), antibody dependent NK cell activation (ADNKA) and FcγR binding were all significantly lower in neuroPASC compared to PASC (**Fig 1B, D**). These data show that individuals who developed neuroPASC had overall lower SARS-CoV-2 specific antibody responses. Whether this diminished systemic SARS-CoV-2 response contributes to incomplete clearance of the virus or simply acts as a marker of a defective overall humoral immune response remains unclear. While emerging data point to overall stability and durability in the natural humoral immune response to SARS-CoV-2,^29^ it is plausible that diverging antibody signatures between groups may be attributable to potential differences in timing from infection in the neuroPASC and no PASC groups. However, based on the onset of the pandemic in Italy, where March 1^st^ 2020 was the earliest potential time of infection in the cohort, the longest potential sampling delay would be a median of 52 days post-infection (IQR 34-125), which is similar to the time of collection for the no PASC group (2-4 months post-infection). Thus, collectively these data point to significant overall differences in the level of systemic SARS-CoV-2 specific humoral immunity in individuals with neuroPASC compared to no PASC.

**Fig 1.**
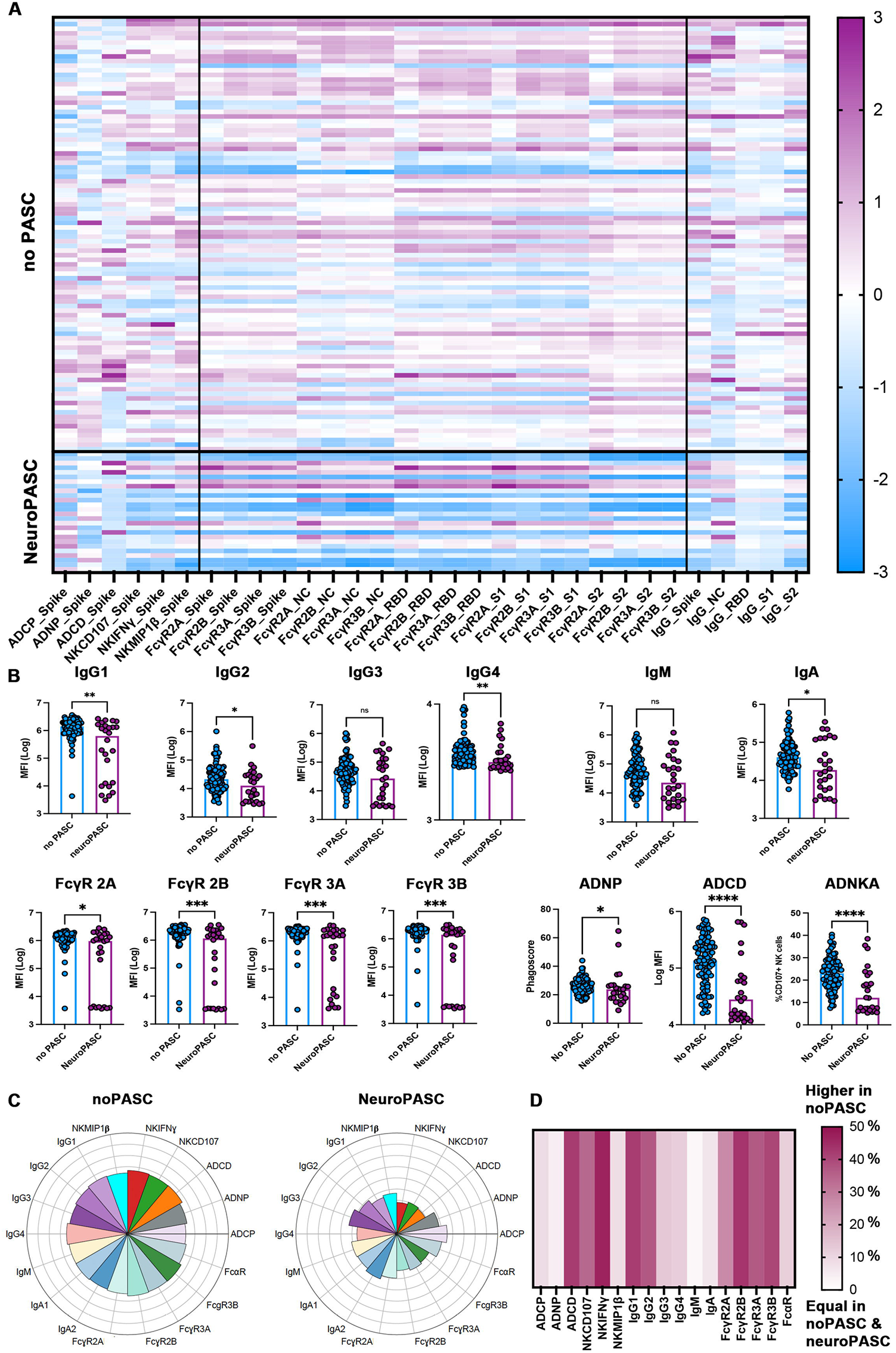
Antibody responses to SARS-CoV-2 differentiate neuroPASC from no PASC. **(A)** Heatmap indicating antibody functions (antibody-dependent complement deposition, ADCD; cellular phagocytosis, ADCP; neutrophil phagocytosis, ADNP; NK activation with production of CD107a, IFNγ and MIP1β), capacity to bind Fcγ receptors (FcγR 2A, 2B, 3A, 3B) and total IgG of different SARS-CoV-2 specific antigens (Spike, S1, S2, RBD and NC) in individuals who did not develop (no PASC) or developed neurological PASC (neuroPASC). Each row corresponds to a single individual. Z-scores, positive (i.e. higher than the mean) in purple, negative (i.e. lower than the mean) in blue. NC= nucleocapsid, RBD=receptor binding domain **(B)** Bar dot plots show the level of IgG subclasses (IgG1, IgG2, IgG3, IgG4), IgM and IgA, FcγR binding capacity (2A, 2B, 3A, 3B) and functions (ADCD, ADNP, ADNKA with production of CD107a) of Spike-specific antibodies in individuals who did not develop (no PASC, in blue) or developed neurological PASC (neuroPASC, in purple). MFI=Median Fluorescence Intensity. Mann-Whitney, *p<0.05, **p<0.01, ***p<0.001 ****p<0.0001. **(C)** Radial plots indicate the levels of IgG subclasses (IgG1, IgG2, IgG3, IgG4), IgM, IgA1, IgA2, binding capacity to FcγR (2A, 2B, 3A, 3B), FcαR, and functions (ADCD, ADCP, ADNP, ADNKA with production of CD107a, IFNγ and MIP1β) of Spike-specific antibodies in individuals who did not develop (noPASC) or developed neurological PASC (neuroPASC). Each sector represents a z-scored antibody feature. **(D)** Heatmap indicating the average difference (%) of Spike-specific functions (ADCD, ADCP, ADNP, ADNKA with production of CD107a, IFNγ and MIP1β), binding capacity to FcγR (2A, 2B, 3A, 3B), FcαR, IgG subclasses (IgG1, IgG2, IgG3, IgG4), IgM and IgA between individuals with no PASC and with neuroPASC. Each column represents an antibody feature. Positive values indicate features enriched in no PASC, null values indicate features equally present in both no PASC and neuroPASC. None of the features were enriched in neuroPASC.

### NeuroPASC is associated with increased antibody responses to common Coronaviruses

Next, we aimed to understand whether lower SARS-CoV-2 specific antibody responses in neuroPASC were uniquely diminished to SARS-CoV-2, or whether individuals with neuroPASC might have deficient humoral responses to other viruses as well. Thus, we profiled responses to Epstein Barr virus (EBV), Influenza virus (Flu), herpes simplex virus (HSV1) and additional common Coronaviruses (229E, OC43, NL63, HKU1). No difference in the antibody responses to EBV, Flu or HSV1 were observed between no PASC and neuroPASC (**Fig 2A, Suppl Fig 1**). However, surprisingly, antibody responses to other common Coronaviruses (in particular 229E), including IgG1 titers and FcγR binding levels, were enriched in individuals with NeuroPASC (**Fig 2B**). These data argue that individuals with neuroPASC generate equivalent responses to common viruses, but exhibit a significant bias towards elevated responses to other non-SARS-CoV-2 Coronaviruses, potentially a marker of imprinting or original antigenic sin.^30^

**Fig 2.**
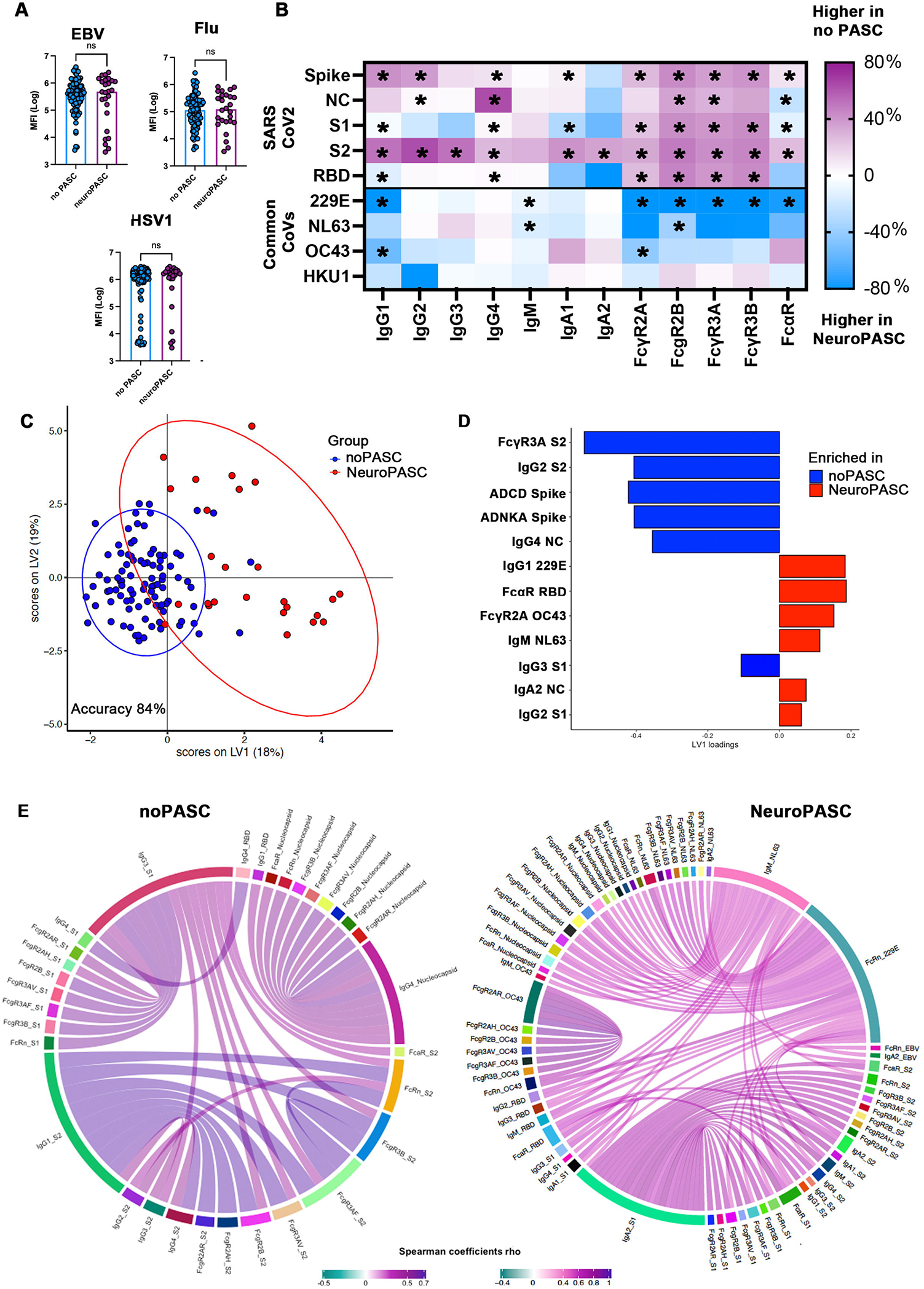
NeuroPASC is characterized by low antibody responses to SARS-CoV-2 and increased responses to common Coronaviruses. **(A)** Bar dot plots show the level of IgG1 antibodies against non-coronaviruses (EBV, Flu, HSV1) in individuals who did not develop (no PASC, in blue) or developed neurological PASC (neuroPASC, in purple). MFI=Median Fluorescence Intensity. Mann-Whitney, ns= statistically not significant. **(B)** Heatmap indicates the average difference (%) of SARS-CoV-2 -specific (Spike, S1, S2, RBD and NC) and common Coronaviruses-specific (229E, OC43, NL63, HKU1) IgG subclasses (IgG1, IgG2, IgG3, IgG4), IgM and IgA1, IgA2, binding capacity to FcγRs (2A, 2B, 3A, 3B), FcαR, between individuals with no PASC and with neuroPASC. Each column represents an antibody feature. Positive values (purple) indicate features enriched in no PASC, negative values (blue) indicate features enriched in neuroPASC, null values (white) indicate features equally present in both no PASC and neuroPASC. Common CoVs = other non-SARS-CoV-2 common Coronaviruses; NC= SARS-CoV-2 nucleocapsid; RBD= SARS-CoV-2 receptor binding domain. Mann-Whitney was used to define statistically significant differences at a univariate level between no PASC and neuroPASC, *p<0.01 or less. (**C-D**) Multivariate analysis of antibody signatures in individuals with neuroPASC and no PASC. Partial least square discriminant analysis (PLSDA) on LASSO-selected features was used to resolve antibody profiles in neuroPASC and no PASC. Dots represent individual samples (no PASC, blue; neuroPASC, red) across SARS-CoV-2 specific (Spike, S1, S2, RBD, NC), and common Coronaviruses (229E, OC43, NL63, HKU1) specific antibody features. Bar graph shows LV1 loadings of LASSO-selected features ranked by their Variable Importance in Projection (VIP). Features enriched in no PASC are in blue, features enriched in neuroPASC are in red. Ten-fold cross validation was performed, resulting in 84% cross validation accuracy (p<0.01). **(E)** Chord plots indicate the Spearman correlation coefficients between the LASSO-selected features enriched in no PASC (left) and in neuroPASC (right), and the non-LASSO selected antibody features across SARS-CoV-2 antigens (Spike, S1, S2, RBD, NC), and other common Coronaviruses antigens (229E, OC43, NL63, HKU1). Only coefficients >0.5 are plotted, p<0.01 after Benjamini-Hochberg correction for multiple comparisons. Positive values represent direct correlations, negative values represent inverse correlations (none of the features were inversely correlated with any other feature).

To further objectively define the differences in the humoral immune response induced in neuroPASC compared to no PASC, we next performed a multivariate analysis using all SARS-CoV-2 and other common Coronaviruses-specific antibody data.^25^ A Least Absolute Shrinkage and Selection Operator (LASSO) was initially used to define the minimal humoral features that separated the two antibody profiles, to avoid statistical overfitting. Using this minimal set of features, a Partial Least-Squares Discriminant Analysis (PLSDA) was then used to visualize the separation between the groups. Robust separation was observed between the NeuroPASC and no PASC antibody profiles (**Fig 2C**, model accuracy 84%, validation p<0.01), with the variation in pathogen-specific antibody profiles across the groups largely driven along latent variable 1 (LV1). Twelve of the total 186 features included in the model were required to separate the 2 groups (**Fig 2D**), with 6 SARS-CoV-2 specific features enriched in individuals with no PASC, and 6 SARS-CoV-2 and common Coronaviruses features enriched in NeuroPASC. Specifically, no PASC antibody profiles targeted several regions of the Spike and the Nucleocapsid antigen, pointing to an overall expanded SARS-CoV-2 response that was highly functional. Conversely, the NeuroPASC response was marked by IgA FcαR-binding antibodies to the Nucleocapsid and the SARS-CoV-2 receptor binding domain (RBD), in addition to an expanded 229E-, OC43- and NL63-specific responses, pointing to an inflammatory mucosal-specific FcαR-biased response in parallel to expanded common Coronaviruses responses.

Moreover, given that the LASSO algorithm minimizes the number of features used in the model, we next aimed to define the additional antibody features that were co-correlated with the most discriminating features, to gain enhanced immunological insights into the antibody profile differences across the groups. Highly divergent co-correlate networks were observed between neuroPASC and no PASC individuals (**Fig 2E**). No PASC subjects displayed strong within SARS-CoV-2 antigens (nucleocapsid, S1, S2) correlations, excluding other viral responses. Conversely, individuals with NeuroPASC exhibited SARS-CoV-2 responses that were highly correlated with common Coronaviruses responses. Of note, the most strongly correlated features in neuroPASC individuals involved nucleocapsid and Spike S2-domain-specific responses, 2 highly conserved regions in both SARS-CoV-2 and common Coronaviruses. Moreover, SARS-CoV-2 specific FcγR-binding responses and broad IgM-responses to NL63 were significantly associated with SARS-CoV-2 specific Nucleocapsid responses. An independent OC43 co-correlate network was observed across FcγR binding levels, pointing to the potential expansion of an independent network of OC43-specific responses with less cross-reactivity to SARS-CoV-2 (**Fig 2E**). Taken together, these data suggest that individuals who do not develop neuroPASC mount a potent and focused immune response to SARS-CoV-2. Conversely, individuals with NeuroPASC evolve a common Coronaviruses focused response, that co-evolves with a dampened SARS-CoV-2 specific response, that may result in partial or compromised control and clearance of the virus.

### CSF is populated by IgG1 and phagocytosis-mediating antibodies

Our data point to a systemic off-target activation of humoral responses towards other common Coronaviruses in the development of neuroPASC, a marker of potential imprinting or original antigenic sin. However, given that these individuals developed signs and symptoms involving the CNS, such as encephalitis with seizures and cognitive deficits, we hypothesized that humoral responses within the brain might directly contribute to CNS dysfunction, and might be compartmentalized. Thus, we next explored the antibody profiles within the CSF from our neuroPASC subjects only. Overall, we observed attenuated CSF SARS-CoV-2 antibody responses (Spike, S1, S2, RBD, nucleocapsid) compared to serum-profiles (**Fig 3A**), including significantly lower Spike-specific titers, FcγR binding capacity and functions (**Fig 3B, C**). Focusing on Spike-specific features, all isotypes and subclasses were detected in the serum, whereas CSF-profiles were characterized by a focused IgG1 (and absence of IgM) response (**Fig 3C, D**). Moreover, the CSF was characterized by a trend towards higher phagocytosis-inducing antibodies (ADCP and ADNP), and exclusion of complement-fixing and NK cell-activating antibodies (**Fig 3B, C, E**). These data show clear compartmentalization of antibodies with specific IgG subclasses and functionality within the CSF. Moreover, because a more diverse isotype/subclass profile would be expected if antibody production occurred within the CSF, the data presented here suggest that SARS-CoV-2 specific antibodies are selectively transferred from the serum into the brain across the blood-brain-barrier and are unlikely to be produced intrathecally (within the CNS).

**Fig 3.**
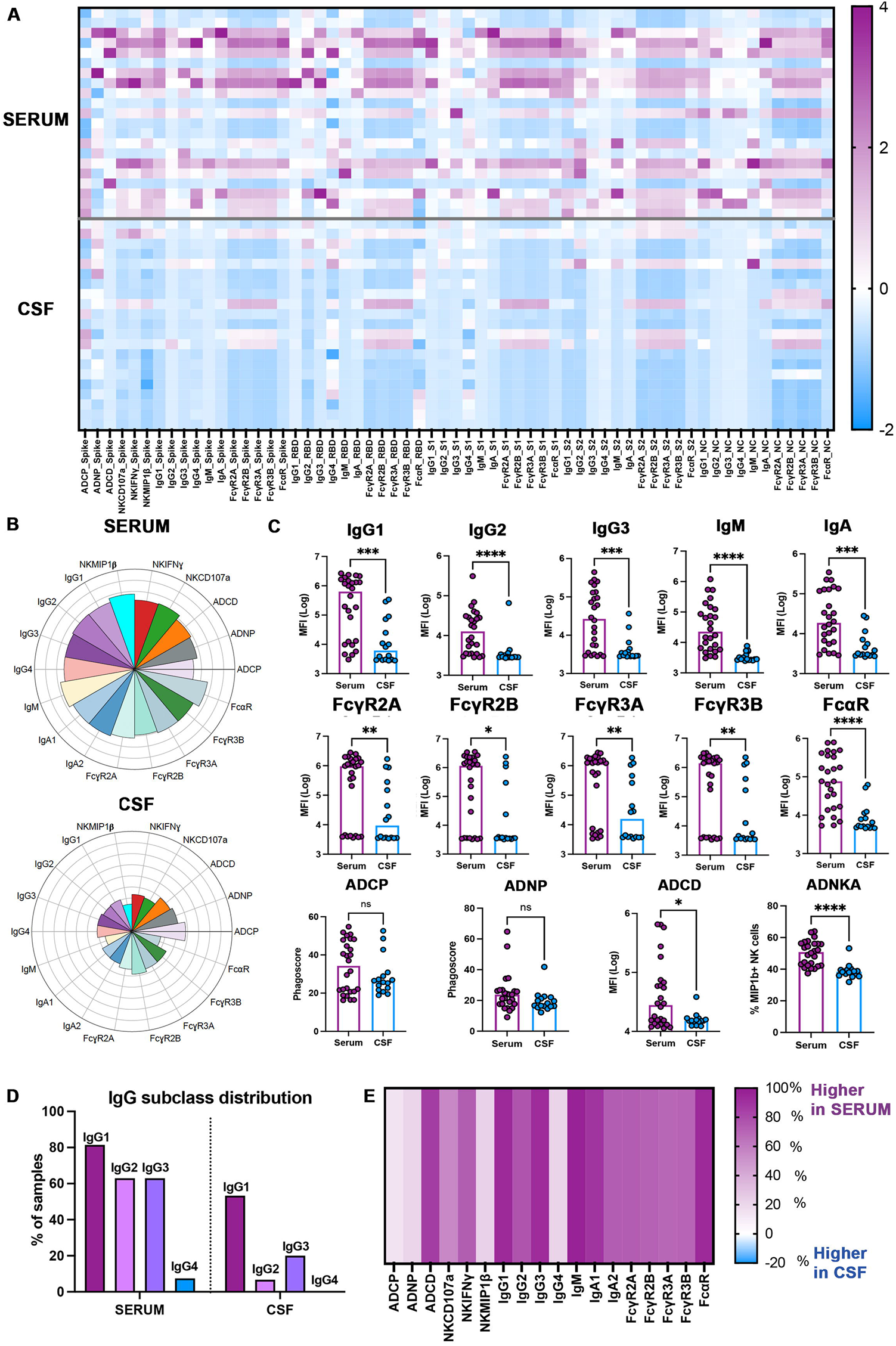
Compartmentalization of antibody responses to SARS-CoV-2 in serum and CSF in individuals with neuroPASC. **(A)** The heatmap indicates functions (ADCD, ADCP, ADNP, ADNKA with production of CD107a, IFNγ and MIP1β), IgG subclasses (IgG1, IgG2, IgG3, IgG4), IgM, IgA, capacity to bind to Fcγ receptors (FcγR2A, 2B, 3A, 3B) and FcαR of antibodies to different SARS-CoV-2 specific antigens (Spike, S1, S2, RBD and NC) in serum and CSF from individuals with neuroPASC. Each row corresponds to a single sample. Z-scores, positive (i.e. higher than the mean) in purple, negative (i.e. lower than the mean) in blue. NC= nucleocapsid, RBD=receptor binding domain **(B)** Radial plots indicate levels of IgG subclasses (IgG1, IgG2, IgG3, IgG4), IgM and IgA1, IgA2, binding capacity to FcγR (2A, 2B, 3A, 3B), FcαR, and functions (ADCD, ADCP, ADNP, ADNKA with production of CD107a, IFNγ and MIP1β) of Spike-specific antibodies in serum and CSF from individuals with neuroPASC. Each sector represents a z-scored antibody feature. **(C)** Bar dot plots show the level of IgG subclasses (IgG1, IgG2, IgG3), IgM and IgA, binding capacity to FcγR (2A, 2B, 3A, 3B) and FcαR, and functions (ADCD, ADNP, ADNKA) of Spike-specific antibodies in serum (purple) and CSF (blue) from individuals with neuroPASC. MFI=Median Fluorescence Intensity. Mann-Whitney, *p<0.05, **p<0.01, ***p<0.001 ****p<0.0001. **(D)** The bar plots indicate the percentage of neuroPASC subjects/samples with positive Spike-specific antibody responses of each IgG subclass in serum and CSF. Positive responses were considered if MFI of each sample was 2-fold higher than the median MFI of PBS level (background). **(E)** Heatmap indicating the average difference (%) of Spike-specific IgG subclasses (IgG1, IgG2, IgG3, IgG4), IgM and IgA1, IgA2, binding capacity to FcγR (2A, 2B, 3A, 3B) and FcαR, between CSF and serum from individuals with neuroPASC. Each column represents an antibody feature. Positive values (purple) indicate features enriched in serum, negative values (blue) indicate features enriched in CSF, null values (white) indicate features equally present in serum and CSF. No features were enriched in CSF.

### Functional coordination in NeuroPASC is higher in serum than in CSF

To mine for differences in the antibody responses between compartments, we next explored the overall humoral architecture against SARS-CoV-2, common Coronaviruses, and additional common infections within the serum and CSF. Interestingly, not only SARS-CoV-2 responses were enriched in the serum compared to the CSF, but also responses to common Coronaviruses and other viruses (EBV, HSV1, Flu) (**Fig 4A**). Likewise, this serum enrichment pertained to all antibody isotypes, subclasses, and FcγR binding levels (**Fig 4A**). Moreover, the humoral immune response was more coordinated in the serum compared to the CSF (**Suppl Fig 2**), likely related to the paucity of humoral immune responses found in the CSF. These data further support a sieving of antibodies from the periphery across the blood-brain barrier, rather than local production of antibodies in the CNS.

**Fig 4.**
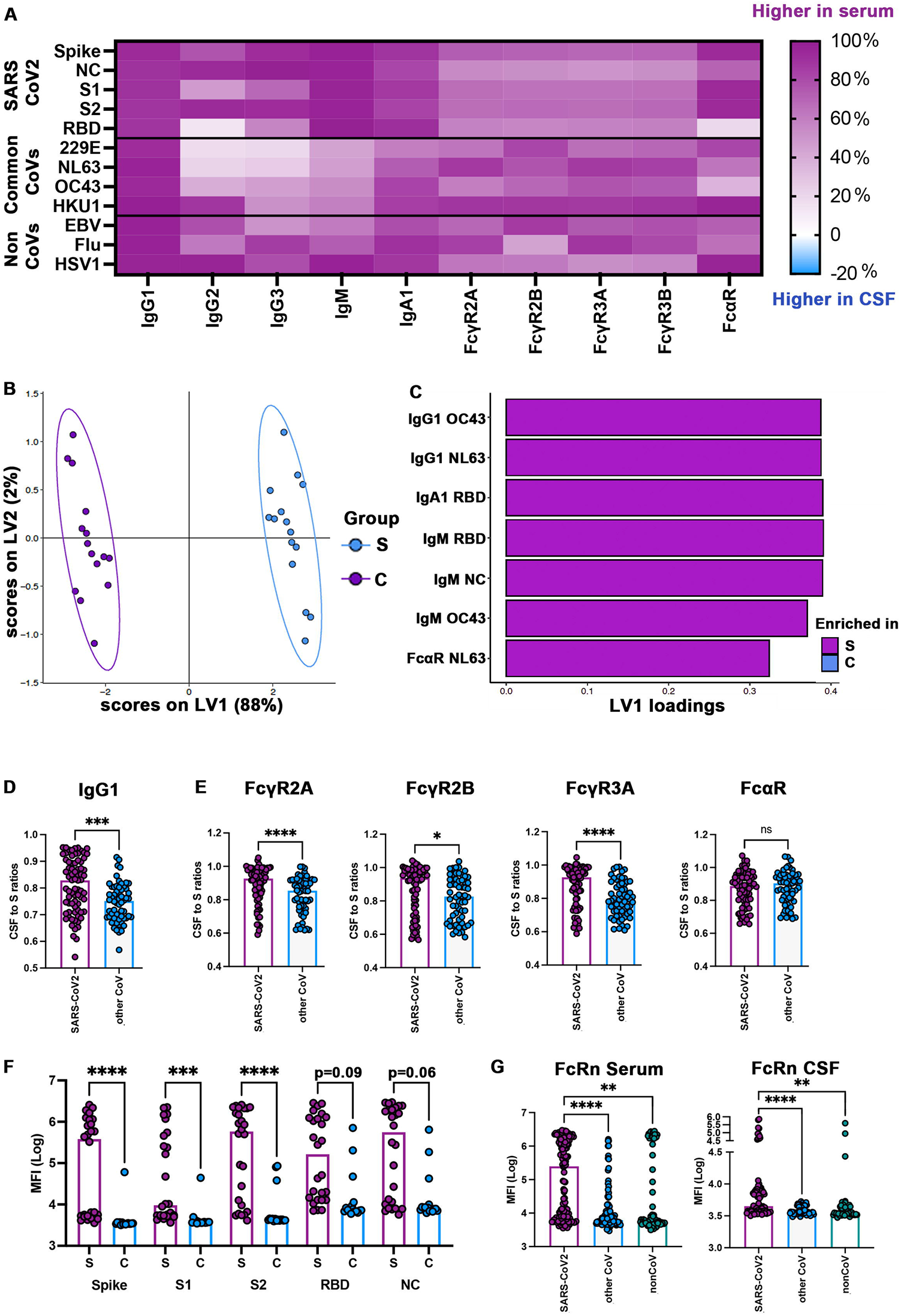
CSF and serum antibody responses to SARS-CoV-2 and other common Coronaviruses in neuroPASC individuals. **(A)** Heatmap indicating the average difference (%) of antibody responses to SARS-CoV-2 (Spike, S1, S2, RBD and NC), other common Coronaviruses (229E, OC43, NL63, HKU1) and non-coronaviruses (EBV, Flu, HHV1), including IgG subclasses (IgG1, IgG2, IgG3, IgG4), IgM and IgA, binding capacity to FcγR (2A, 2B, 3A, 3B) and FcαR, between serum and CSF from individuals with neuroPASC. Positive values (purple) indicate features enriched in serum, negative values (blue) indicate features enriched in CSF, null values (white) indicate features equally present in both serum and CSF. There were no features enriched in CSF. Common CoVs = other non-SARS-CoV-2 common Coronaviruses (229E, OC43, NL63, HKU1); NC= nucleocapsid; RBD= receptor binding domain **(B-C)** Multivariate analyses of antibody signatures in serum and CSF from individuals with neuroPASC. Multilevel partial least square discriminant analysis (M-PLSDA) on LASSO-selected features was used to resolve antibody profiles in serum:CSF pairs. Dots represent individual samples (serum, blue; CSF, purple) across SARS-CoV-2 antigens (Spike, S1, S2, RBD, NC), and common Coronaviruses (229E, OC43, NL63, HKU1). Bar graph shows LV1 loadings of LASSO-selected features ranked by their Variable Importance in Projection (VIP). Features enriched in serum are in purple, no features were enriched in CSF. Ten-fold cross validation was performed, resulting in 100% cross validation accuracy (p<0.01). **(D-E)** Bar dot plots indicate CSF to serum ratios of IgG1, FcγR (2A, 2B, 3A) and FcαR binding capacity for SARS-CoV-2 (Spike, S1, S2, RBD, NC antigens combined) and other common Coronaviruses (229E, OC43, NL63, HKU1 combined) antibodies in individuals with neuroPASC. Mann-Whitney, *p<0.05, ***p<0.001 ****p<0.0001. CoV=common Coronaviruses; NC= nucleocapsid; RBD= receptor binding domain **(F)** Bar dot plots show the binding capacity of antibodies to the neonatal Fc receptor (FcRn) of serum and CSF SARS-CoV-2 antibodies targeting different antigens (Spike, S1, S2, RDB, NC) in individuals with neuroPASC. Mann-Whitney, *p<0.05, **p<0.01, ***p<0.001 ****p<0.0001. MFI=Median Fluorescence Intensity; NC=nucleocapsid; RBD=receptor binding domain **(G)** Bar dot plots indicate the FcRn binding capacity of serum and CSF SARS-CoV-2 antibodies (Spike, S1, S2, RBD, NC, combined), compared to common Coronaviruses (229E, OC43, NL63, HKU1 combined) and non-coronaviruses (EBV, Flu, HSV1 combined). Kruskal Wallis test, Dunn’s correction for multiple comparisons. **p<0.01, ****p<0.0001. MFI=Median Fluorescence Intensity; NC= nucleocapsid; RBD= receptor binding domain

To further probe differences across compartments, we performed a LASSO/M-PLSDA multivariate analysis using all SARS-CoV-2 and common Coronaviruses antibody data (**Fig 4B,C**). Striking separation was observed in the antibody profiles between the serum and the CSF (**Fig 4B**, model accuracy 100%, validation p<0.01), driven by as few as 7 of the 141 analyzed features, including both SARS-CoV-2 and common Coronaviruses antibody IgG, IgA, and IgM responses, which were all enriched in the serum (**Fig 4C**).

To next define the relationship between the CSF and the serum humoral immune response we next analyzed the CSF:serum ratios of antibodies to SARS-CoV-2 (all antigenic targets combined) and other common Coronaviruses (229E, OC43, NL63, HKU1 combined). Interestingly, we observed higher CSF:serum ratios for SARS-CoV-2 specific antibodies, including elevated titers (IgG1) and FcγR (but not FcαR) binding capacity (**Fig 4D,E**), pointing to a clear selective enrichment of IgG-dominated SARS-CoV-2 specific, but not other pathogen-specific, antibody immunity in the CSF. Furthermore, serum SARS-CoV-2 specific antibodies (in particular those targeting Spike and S2) exhibited strong FcRn binding capacity compared to weaker binding by CSF antibodies (**Fig 4F**). Also, FcRn binding by serum SARS-CoV-2 antibodies was significantly higher than binding by other non-SARS-CoV-2 antibodies, whereas this difference was less pronounced in the CSF (**Fig 4G**), suggesting enhanced FcRn-mediated transfer of SARS-CoV-2 specific antibodies from the circulation to the CSF with decreased recycling from the CSF to the circulation.

Antibody transfer to the brain is thought to occur via the neonatal Fc-receptor (FcRn), which should transfer equal amounts of all IgG antibodies into the CSF.^31^ However, the preferential transfer of SARS-CoV-2 specific immunity pointed to a further selection of particular antibody subpopulations from the blood into the brain. Moreover, differences of CSF:serum ratios between SARS-CoV-2 and other Coronaviruses antibodies were more pronounced for activating FcγR binding (FcγR2A and FcγR3A) compared to inhibitory FcγR2B (**Fig 4E**), pointing to a potential selective role of activating:inhibitory receptors in transfer across the compartments. Overall, our results suggest selective/preferential transfer of SARS-CoV-2 specific antibodies (but not antibodies to other viruses) from the circulation to the CSF, driven in a FcγR-dependent manner in collaboration with FcRn. These data point to a clear selective transfer of antibodies to specific pathogens to the brain likely related to the co-selection of recently generated, potentially more inflammatory antibodies, to fight infection in the brain.

### Antibody profiles correlate with NeuroPASC outcome

We next aimed to understand whether specific antibody profiles might track with differential clinical outcome in individuals with neuroPASC. Therefore, neuroPASC individuals were classified as having *good* (mRS < 2) or *poor* clinical outcome (mRS ≥ 2) at last follow-up. Interestingly, individuals with good outcome exhibited higher serum SARS-CoV-2 specific IgG1 titers and FcγR binding capacity, whereas those with poor outcome showed increased antibody responses to other common Coronaviruses (229E, OC43, NL63) (**Fig 5A**).

**Fig 5.**
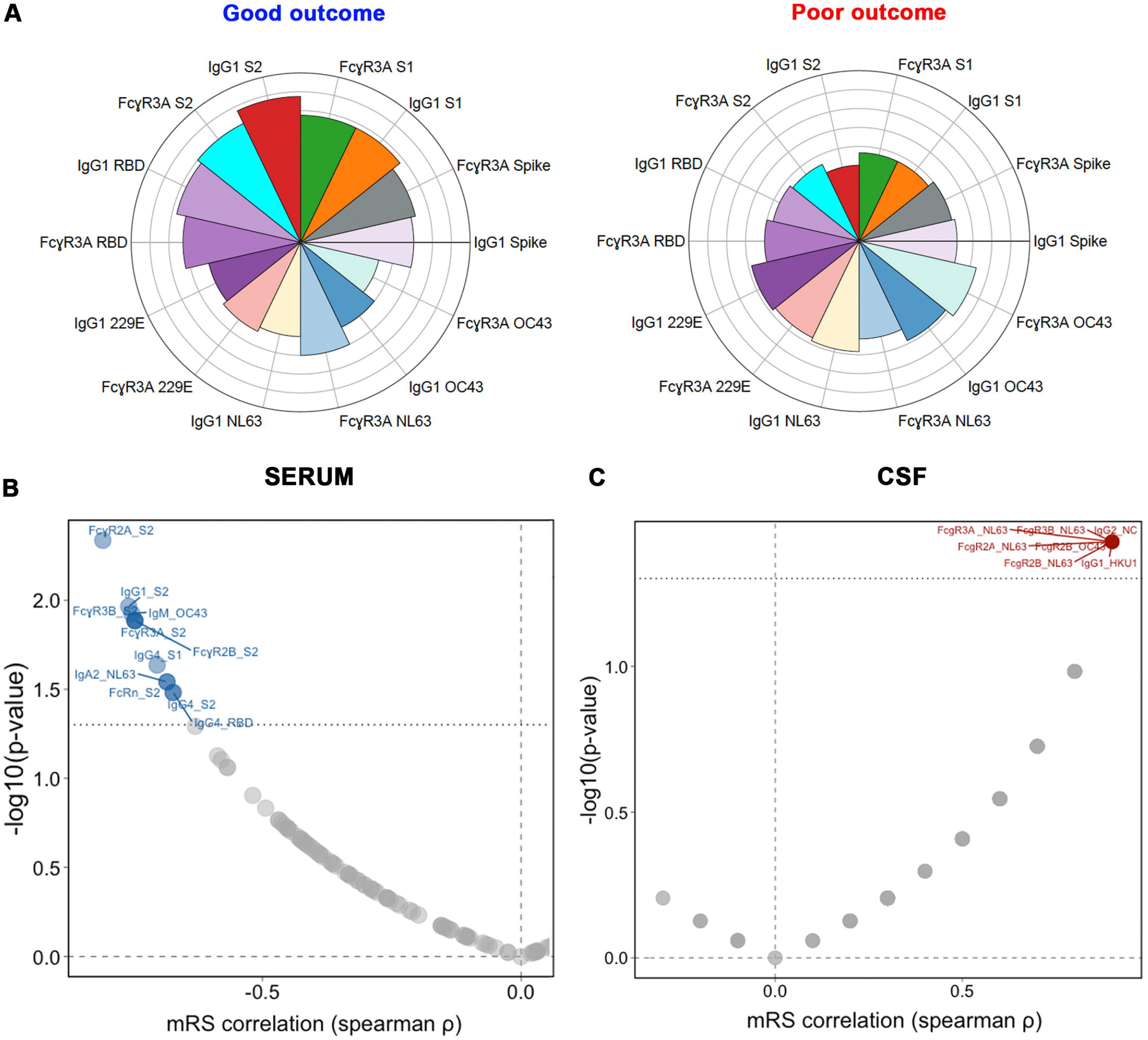
CSF and serum antibody signatures associated with good versus poor outcome in neuroPASC individuals. **(A)** The radial plots indicate the levels of IgG1 and FcγR2A binding capacity of SARS-CoV-2 antibodies (Spike, S1, S2, RBD, upper half circle) and common Coronaviruses (229E, OC43, NL63, lower half circle) in individuals with neuroPASC, divided into those with good versus poor outcome. Each sector represents a z-scored antibody feature. RBD=receptor binding domain **(B-C)** Volcano plots showing the correlation of each SARS-CoV-2 and common Coronaviruses (229E, OC43, NL63, HKU1)-antibody feature with modified Rankin Scale (mRS). Spearman correlation coefficients are indicated in the x-axis (positive correlation on the right, red; negative correlation on the left, blue), and the statistical significance is indicated in the y-axis (-log10[p-values]). Values above black dashed line indicate statistically significant correlations (p adjusted value<0.01, Benjamini-Hochberg correction for multiple comparisons). Negative correlations, indicating good outcome, were identified with serum antibodies (left graph, in blue), whereas negative correlations, indicating poor outcome, were identified with CSF antibody profiles (right graph, in red).

Finally, we aimed to explore whether the clinical outcome in neuroPASC individuals correlated with antibody features in serum and CSF. Interestingly, we observed that serum SARS-CoV-2 specific antibody features (in particular S2-specific IgG1, FcγR2A, FcγR2B, FcγR3A, FcγR3B binding capacity) negatively correlated with mRS (**Fig 5B**), indicating that stronger serum SARS-CoV-2 responses are clearly linked to better outcome in neuroPASC individuals, likely through improved clearance or control of the infection. In contrast, common Coronaviruses features, including OC43-specific FcγR2A, NL63-specific FcγR2A, 2B, 3A, 3B binding and HKU1-specific IgG1, dominated the CSF-antibody profiles that all positively correlated with mRS (**Fig 5C**), indicating a relation between non-SARS-CoV-2 antibody features and worse outcome. These results suggest a role for peripheral SARS-CoV-2 antibodies as biomarkers of favorable outcome in neuroPASC individuals, and expanded CSF common Coronaviruses antibody responses as markers of poor clinical profiles. Collectively, these data point to the potential critical importance of original antigenic sin or imprinting in shaping the humoral immune response to SARS-CoV-2 immunity, representing, particularly in the CSF, a key prognostic biomarker of neuroPASC disease.

### Data availability

Data will be available to qualified researchers upon reasonable request

## DISCUSSION

Post-acute neurological complications (neuroPASC) occur in a significant portion of the patients with SARS-CoV-2 infection. These symptoms often occur in previously healthy individuals, even in the setting of asymptomatic or mild disease, and can persist for several months with significant impact on functional and work capacity.^32^ However, both biomarkers and mechanisms that explain these debilitating secondary effects of SARS-CoV-2 infection remain ill defined, but could revolutionize patient care and the development of disease-modifying therapies.

Emerging studies in individuals with PASC have begun to collectively point to anomalous immune mechanisms, marked by perturbated inflammatory cytokine profiles,^33-35^ altered cellular transcriptional signatures,^36,37^ and auto-antibodies production.^36,38,39^ However, given the heterogeneity of PASC clinical manifestations, including individuals with neurologic, respiratory, vascular, or rheumatologic complications, it is possible that unique pathways may be triggered during SARS-CoV-2 infection that may lead to distinct PASC phenotypes. In the setting of neuroPASC, new data point towards immune-mediated mechanisms as a driver of disease. Thus, here we applied a systems serology approach and identified distinctive antibody signatures in neuroPASC individuals, characterized by compromised SARS-CoV-2 specific humoral responses in association with enhanced common Coronaviruses immunity. Moreover, these common Coronaviruses immune responses were likely transferred from the circulation into the CSF, rather than generated within the CNS, and were directly associated with neuroPASC outcome. These data, collectively, point to a potentially incomplete maturation of the SARS-CoV-2 response, biased by a pre-existing humoral immunity to common Coronaviruses, that may lead to development of neuroPASC due to partial or delayed viral clearance and persistent neuroinflammation.

While it has been speculated that a persistent reservoir of SARS-CoV-2 in the CNS could contribute to neuronal damage in neuroPASC, analyses of CSF from living individuals^7,40^ as well as autopsy studies,^41^ have rarely detected active viral replication in the brain. Moreover, viral replication in the CNS would likely be accompanied by recruitment of B cells in the brain, that would convert to antibody-secreting plasma cells capable to switch and locally generate large numbers of polyclonal antibodies of various Ig subclasses, in response to the local presence of antigen. Instead, individuals with neuroPASC had highly compartmentalized CSF-antibody profiles, characterized by lower levels, but highly focused IgG1 phagocytic profiles. This is in line with previous observations in patients with acute COVID-19 and neurological manifestations, reporting CSF SARS-CoV-2 specific antibodies, whose target epitopes were different from serum antibodies,^42^ and were associated with compartmentalized cytokine production.^43^ While CSF-specific antibody profiles that we observed in neuroPASC were also present in the systemic circulation, the systemic response was much more diverse, arguing for the selective transfer of antibodies into the CNS. In homeostatic conditions, antibodies are thought to circulate into the brain through an FcRn-mediated transcytosis mechanism across the blood-brain barrier.^44,45^ FcRn binds to IgG1 with higher affinity than to other antibody subclasses, but additional selectivity was observed across antigen-specificity and FcγR binding capacities, arguing for the additional role of activating, but not inhibitory, FcγRs as co-transporters of antibodies into the CNS, potentially as a shield against any virus that should breach the blood-brain-barrier. Similarly to our previous reports on HIV,^46^ these data suggest the presence of novel antibody transport mechanisms into the brain, but also point to limited immunologic evidence of persistent active viral replication, and antigen generation, into the brain.

The role of dysregulated immune mechanisms in the development of neurological complications of SARS-CoV-2 infection has become increasingly accepted. Most studies have focused on neurologic manifestations during the acute phase of COVID-19, and have shown the enrichment of both systemic immune activation^47-49^ and CSF-specific increased cytokine levels.^50,51^ Moreover, studies have shown evidence of higher levels of SARS-CoV-2 specific antibodies, especially in the CSF, in individuals with more severe neurological disease and CNS damage.^43,52-53^ These data point towards a role for a hyperinflammatory state and neuroinflammation in the development of neurological manifestations during acute SARS-CoV-2 infection.^51^ However, the role of humoral and other immune responses in the persistence (or delayed onset) of neurological symptoms beyond acute COVID-19 disease is yet to be clarified. Our data show that individuals with neuroPASC exhibited a biased immune response across coronaviruses, with an expanded common Coronaviruses response (in particular 229E, NL63 and OC43) at the expense of an attenuated SARS-CoV-2 response in the serum and CSF. This suggests a mechanism of original antigenic sin, in which previous immune responses to common Coronaviruses might shape subsequent responses to other related viruses, such as SARS-CoV-2, which share high epitope homology. This mechanism, also known as immunologic imprinting, has been observed also for other pathogens, such as influenza virus^30^ and more recently HIV.^54^ These studies demonstrated that, during secondary exposure to pathogens, circulating antibodies produced during the primary humoral response modulate naïve B cell recruitment, blocking or skewing the maturation of the humoral response to the new pathogen. Likewise, in our study we observed a bias towards common Coronaviruses-specific immunity as a marker of neuroPASC, that may inhibit the evolution of new B cell responses to SARS-CoV-2, that may ultimately lead to decreased or delayed viral clearance. Interestingly, this antibody signature was amplified in individuals with poor clinical outcome, across both compartments. These data suggest a previously unappreciated role for immunologic imprinting from other Coronaviruses that may induce a defective antibody-mediated control of SARS-CoV-2 infection, resulting in incomplete or delayed virus clearance, and persistence of systemic immune activation and neuroinflammation involved in the pathogenesis of neuroPASC. Whether the markers identified here are simply biomarkers or mechanistic players in neuroPASC remains unknown, but may support the more effective identification, management, and potential treatment of individuals suffering from neuroPASC.

## Data Availability

All data produced in the present study are available upon reasonable request to the authors

## AUTHORS CONTRIBUTIONS

Collected clinical data and samples: AD, SB, VC, SF, SM. Conceived and designed the experiments: MS, GA. Performed the experiments: MS, NN, DY. Analyzed the data: MS, NN, WJ, YY, DAL. Contributed to interpretation of data: MS, NN, GA. Wrote the paper: MS, GA. Revised the paper for intellectual content: MS, NN, YD, WJ, DY, AD, SB, VC, SF, DAL, SM, GA

## SUPPLEMENTAL FIGURES LEGEND

**Fig 1. Antibody responses to non-coronaviruses are similar between neuroPASC and no PASC**

The heatmap indicates IgG subclasses (IgG1, IgG2, IgG3, IgG4), IgM, IgA1, IgA2, capacity to bind Fcg receptors (2AR, 2AH, 2B, 3AV, 3AF, 3B) and FcαR of EBV-, Flu-, and HSV1-specific antibodies in individuals who did not develop (no PASC) or developed neurological PASC (neuroPASC). Each row corresponds to a single individual. Z-scores, positive (i.e. higher than the mean) in purple, negative (i.e. lower than the mean) in blue. EBV= Epstein-Bar virus; Flu= influenza virus A; HSV1= herpes simplex virus 1.

**Fig 2. Functional coordination in serum and CSF of neuroPASC individuals**

The correlation matrix shows the Spike-specific functions (ADCD, ADCP, ADNP, NK CD107, NK IFNγ, NK MIP1β), IgG subclasses (IgG1, IgG2, IgG3, IgG4), IgM, IgA1, IgA2, and Fc receptor binding (FcγR2A, FcγR2B, FcγR3A, FcγR3B, FcRn, FcαR) in serum and CSF in neuroPASC individuals. Correlation strength is proportional to color intensity (r from -1 to 0 = negative correlation, blue; from 0 to 1 = positive correlation, purple). (Z-scores, Spearman r test, Benjamini-Hochberg correction for multiple comparisons, *p<0.05, **p<0.01, ***p<0.001).

