## Supplementary figures and images for "Neurologic sequalae of COVID-19 are determined by immunologic imprinting from previous Coronaviruses"

### Suppl Fig 1

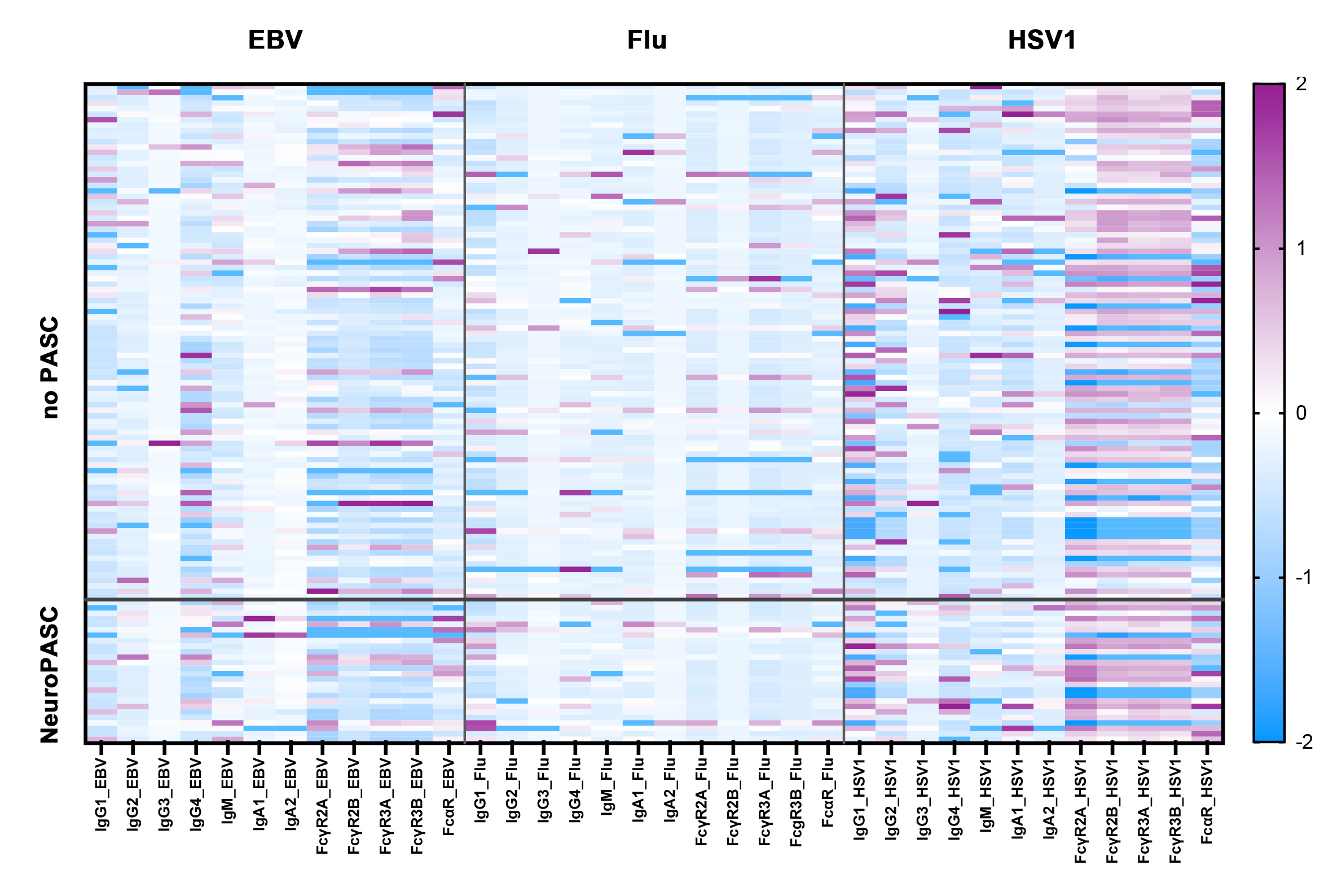

### Suppl Fig 2

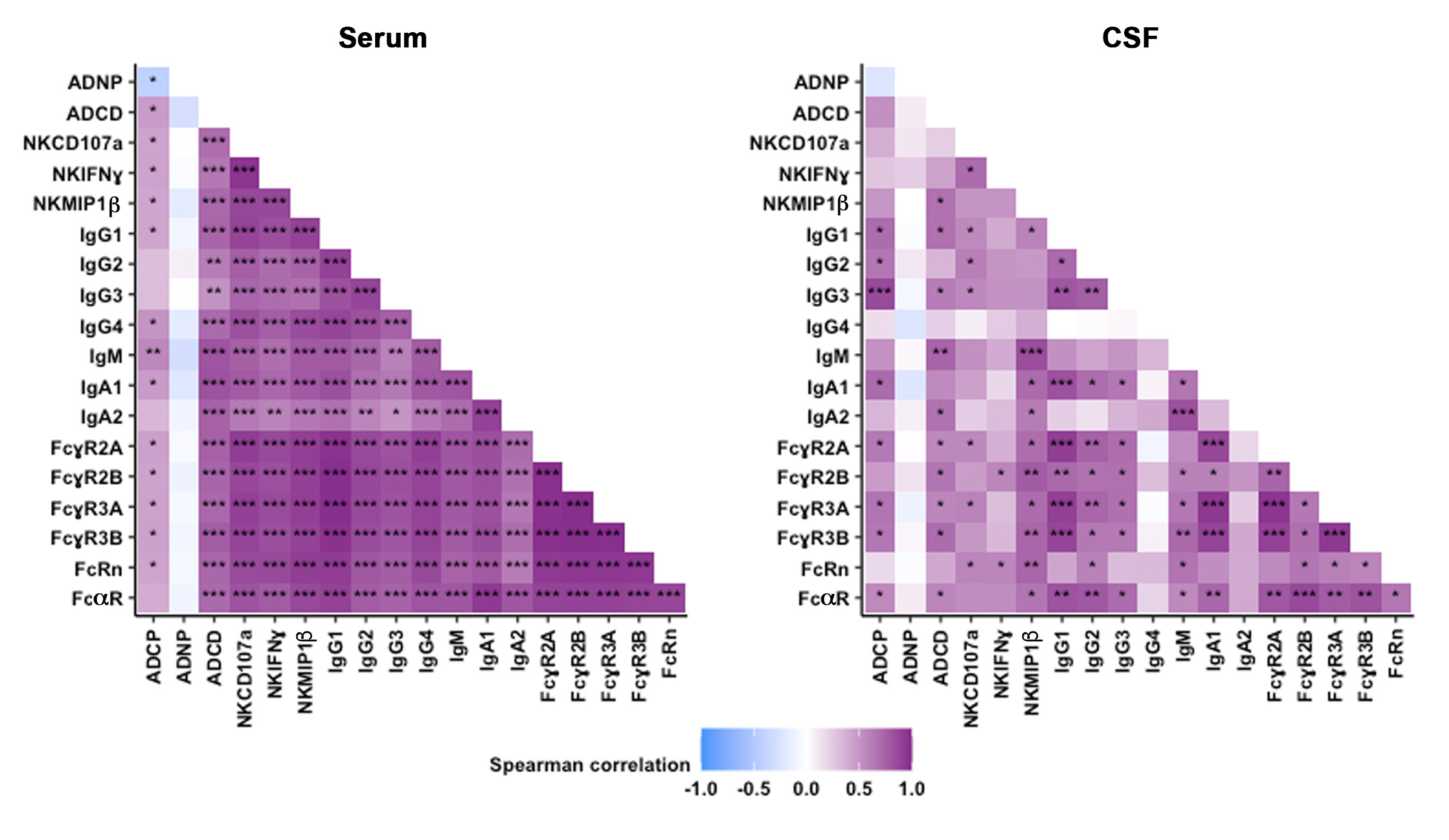
